# Impaired peripheral mononuclear cell metabolism in patients at risk of developing sepsis: A cohort study

**DOI:** 10.1101/2020.12.10.20244707

**Authors:** Velma Herwanto, Ya Wang, Maryam Shojaei, Alamgir Khan, Kevin Lai, Amith Shetty, Stephen Huang, Tracy Chew, Sally Teoh, Marek Nalos, Mandira Chakraborty, Anthony S. McLean, Benjamin MP. Tang

## Abstract

**Purpose:** Dysregulated immune response is a key driver of disease progression in sepsis and known to be associated with impaired cellular metabolism. This association has been studied mostly in the late stage sepsis patients. Here, we investigate whether such impairment in cellular metabolism is present in uncomplicated infection patients who do not develop sepsis.

**Methods:** Forty sepsis (fulfilled Sepsis-3 criteria) and 27 uncomplicated infection patients were recruited from the emergency department along with 20 healthy volunteers. Whole blood was collected for measurement of gene expression, cytokine levels and cellular metabolic functions (including mitochondrial respiration, oxidative stress and apoptosis).

**Results:** Our analysis revealed the impairment of mitochondrial respiration in uncomplicated infection and sepsis patients (p value <0.05), with greater degree of impairment noted in the established sepsis. The impairment was significantly correlated with increased mitochondrial oxidative stress level; the latter was increased in uncomplicated infection and more so in established sepsis patients. Further analysis revealed that the oxidative stress level correlated significantly with cytokine level (tumor necrosis factor-α) and gene expression levels (CYCS, TP53, SLC24A24 and TSPO).

**Conclusions:** These findings suggest that impaired immune cell metabolism is present in infection patients without presenting sepsis, thereby opening potential window for early diagnosis and intervention (e.g. antioxidant therapy) in such patients.

## Introduction

Sepsis is one of the most significant disease burdens in the world (1). A better understanding of sepsis disease mechanism is urgently needed to facilitate development of new therapies. Several pathophysiological mechanisms are involved, including endothelial dysfunction, coagulopathy and dysregulated immune response (2). Among these, dysregulated immune response is thought to play the most critical role in sepsis (3). The dysregulated immune response in sepsis is characterized by impaired innate and adaptive immune responses; both of which have been shown to strongly correlate with poor patient outcomes (4, 5).

It has been demonstrated that impaired cellular metabolism contributes to the dysregulated immune response in sepsis. The impaired cellular metabolism includes reduced cellular oxygen utilization and reduced ATP production (6–8). A number of studies in sepsis patients have confirmed that these impairments are associated with increased organ failures and reduced survival (9, 10).

Recent studies sought to identify defective pathways that are associated with impaired cellular metabolism in peripheral blood mononuclear cell (PBMC) of sepsis patients (11, 12). These studies have identified several defective cellular pathways such as inhibited mitochondrial complex activity and oxygen consumption, and reduced ATP production across different sepsis populations (13, 14). However, these studies share a common limitation; they were conducted in patients with established sepsis or in the late stage of sepsis. Therefore, it is uncertain whether impaired cellular metabolism is present prior to the development of sepsis.

In this study, we sought to measure cellular metabolic state of PBMC in infection phase by studying peripheral blood samples from patients who present to emergency department with suspected infections, in whom sepsis is yet to develop. The overall goal of the study is to detect early changes in the metabolic profile of circulating immune cells in the infection patients and compare them to that of patients who later develop sepsis.

## Material and methods

### Subjects recruitment

This study was approved by Human Research Ethics Committee at our institution (reference number HREC/18/WMEAD/67). Patients were recruited from the emergency department (ED) at Westmead Hospital, New South Wales, Australia, between February 2018 and July 2019. Written informed consent was obtained from all subjects recruited. Inclusion criteria: subjects were eligible if they aged 18 years or older; presented to the ED within the last 24 hours with infection, defined as either (1) positive pathogen identification in any body fluids sampled for microbiological culture, or (2) a suspicion of infection (as determined by treating physician) and received antibiotics. Exclusion criteria: (1) decision not to actively treat or resuscitate the patient at admission; and (2) inability to consent the patient.

### Case definitions

The study participants were assigned into sepsis or uncomplicated infection groups, based on their Sequential Organ Failure Assessment (SOFA) score on admission (≥2 and <2, respectively) in accordance with the international consensus definition of sepsis (“Sepsis-3”) (15, 16). Healthy volunteers who did not have infection or any pre-existing comorbidities were recruited as controls.

### Gene expression profiling

#### RNA extraction

Two and a half millilitres of whole blood, which was collected into PAXgene Blood RNA tube (BD Biosciences), was allowed to stand at room temperature for 2 hours, followed by storage at −20°C for 24 hours. Thereafter, the tubes were transferred to −80°C for long term storage. Total RNA was extracted using PAXgene Blood RNA Kit (QIAGEN) following the manufacturer’s protocol. Quantity and integrity of purified RNA were determined by the RNA integrity number (RIN) using TapeStation (Agilent Technologies Inc.).

### Gene expression by NanoString^⍰^Technology

Gene expression was measured using Nanostring nCounter® Elements™ TagSet (NanoString^⍰^ Technologies Inc) preselected for mitochondrial biogenesis and function panel. The panel includes 90 human genes and 6 housekeeping genes (Table S1). One hundred nanograms of RNA from samples with RIN ≥7.0 was used for setting up the hybridization reaction. A standard nCounter Prep Station was set up on the workstation for post-hybridization processing. After which, sample cartridge was removed, sealed and loaded into the Digital Analyzer to be scanned. Data resolution was set to high (280 images per sample).

### Gene expression data analyses

nCounter data files (RCC files) were imported into the nSolver™ analysis software v4.0 (NanoString Technologies) for quality control and raw mRNA abundance frequencies analysis. Quality control was performed with the built-in negative and positive controls following recommendations by NanoString Technologies. Raw data was normalized against the geometric mean of 6 housekeeping genes included in the codeset. The expression level was presented as fractional difference. It was calculated by dividing the difference between the pooled expression values of patient and of healthy control, with pooled expression values of healthy control. Pathway analysis was performed with Reactome Pathway Portal version 3.2 (17).

### Measurement of serum cytokines

Whole blood collected in tube without anticoagulant was allowed to clot for 30 minutes to 1 hour at room temperature before centrifugation at 1,600 x g for 15 minutes at room temperature. Supernatant was collected and stored at −80°C for further analyses.

Concentrations of cytokines were measured using the Bio-Plex Pro™ □ Human Cytokine Screening 6-Plex Panel (Bio-Rad Laboratories) according to the manufacturer’s protocol. Analytes measured were interleukin 1 beta (IL-1β), interleukin 4 (IL-4), interleukin 6 (IL-6), interleukin 10 (IL-10), tumour necrosis factor alpha (TNF-*α*) and interferon gamma (IFN-γ). Briefly, thawed serum samples were centrifuged at 10,000 x g for 10 minutes at 4°C to remove cells. Fifty microliters of the duplicated serial-diluted standards and serum samples were added to 96-well plate which contained the pre-washed antibody coated microbeads. The plate was sealed, followed by incubation at room temperature for 1 hour with shaking at 850 rotations per minute under dark. The plate was then washed 3 times with Bio-Plex Pro II magnetic plate washer and incubated with detection antibody for 30 minutes followed by 3 times washes and incubation with streptavidin-PE for 10 minutes at room temperature under dark. All the liquids were delivered to the assay plate using a robotic platform (epMotion 5075, Eppendorf). The plate was read with the Bio-Plex 200 system using the Bio-Plex manager v6.1 software.

### Measurement of cellular metabolism

Agilent Seahorse XF Analyser was used to measure two key parameters: oxygen consumption rate (OCR) and extracellular acidification rate (ECAR). These parameters reflect effectiveness of mitochondrial respiration (OCR) and glycolysis (ECAR), which provides an alternative energy pathway to mitochondrial respiration.

#### Sample collection and preparation

Whole blood sample was collected from healthy controls or study participants into tubes containing anticoagulant EDTA. Peripheral blood mononuclear cells were isolated from the whole blood using Ficoll. Isolated PBMCs were cryopreserved in freezing medium (fetal bovine serum (FBS) with 10% DMSO) and stored in Vapor Phase liquid nitrogen tank until further analyses.

#### Cellular metabolism assay

On the day of the assay, PBMCs were thawed and washed in RPMI + 10% FBS, and finally resuspended in Seahorse XF Assay Medium and seeded onto microplate pre-coated with Corning^⍰^Cell-Tak Cell and Tissue Adhesive (Bio-Strategy Pty Ltd) at 5 × 10^5^ cells per well. Each sample was analysed at least in triplicate. Metabolic profiling of PBMC was assessed with the use of cell mito stress test kit (Agilent Technologies, Inc.), which includes three drugs – oligomycin, carbonyl cyanide-4 (trifluoromethoxy) phenylhydrazone (FCCP) and rotenone/ antimycin A. These drugs were pre-titrated and 2 µM oligomycin, 2 µM FCCP and 0.5 µM rotenone/antimycin A were used for our assays. Sequential injections of these drugs allowed the calculations of parameters related to mitochondrial respiration, including basal respiration, maximal respiration, spare respiratory capacity and ATP production (18).

Data was analysed using the Wave Desktop software version 2.6. To normalize the data with cell number, total protein amount was quantified at the completion of the XF assay using Bradford protein assay kit (Bio-Rad) after cells in the wells were lysed with lysis buffer (10 mM Tris, pH 7.4, 0.1% Triton X-100) (19).

## Measurement of Oxidative Stress

### Total cellular reactive oxygen species (ROS)

To detect total cellular ROS, cells were stained with 2’,7’ – dichlorofluorescin diacetate (DCFDA) Cellular ROS Detection Assay Kit (abcam) according to manufacturer’s protocol. Briefly, thawed PBMCs, resuspended in RPMI + 10% FBS, were allowed to rest for 2 hours before staining with 20 µM DCFDA for 30 minutes in 37□C CO_2_ incubator. Stained cells were subjected to analyses by BD FACs CantoII flow cytometer (BD Biosciences). Application setting was applied each time before analyses. Data was analysed using FlowJo v10 (BD Biosciences). Results were expressed as median fluorescence intensity (MFI) after subtracting background from unstained samples.

### Mitochondrial superoxide

To quantify mitochondrial superoxide, cells were stained with MitoSOX™ □ Red (Invitrogen) according to manufacturer’s protocol. Briefly, thawed PBMCs were washed and resuspended in Hank’s Balanced Salt Solution with calcium and magnesium (HBSS/Ca/Mg), followed by staining with 5 µM MitoSOX™□ Red for 10 minutes in 37□C CO_2_ incubator. After staining, cells were washed twice and resuspended in HBSS/Ca/Mg, which were then subjected to flow cytometry analyses as above.

## Measurement of apoptosis

Cell apoptosis was quantified using Annexin V-FITC Apoptosis Detection Kit (abcam) according to manufacturer’s protocol. Cells were washed and resuspended in 1x binding buffer, followed by staining with Annexin V-FITC and propidium iodide (PI). Stained cells were subjected to flow cytometry analyses as above. The Annexin V positive and PI negative cells were assigned as apoptotic cells.

## Statistical analysis and data visualization

All statistical analyses were performed using SPSS version 25 (IBM) and GraphPad Prism 8. For the normally distributed data, analyses were conducted using parametric test. For data that was not normally distributed, data transformation was performed and was then subjected to parametric test. If the transformation was not successful to normalise the data, the non-parametric test was used. Graphs were generated using GraphPad Prism 8. Heatmap was created using Morpheus software from Broad Institute (https://software.broadinstitute.org/morpheus).

## Results

### Patients characteristics

Demographic and clinical characteristics of patients are summarized in Table 1. There was no significant difference in baseline characteristics between uncomplicated infection and sepsis groups except for age (p = 0.002) and number of patients with existing comorbidities (p = 0.015). The average age of sepsis group was higher than that of uncomplicated infection group (means 69 vs 56). Twenty healthy samples were recruited as controls, twelve (60%) of them were male. Their median age was 59 (28 – 72) years old. Sepsis patients had higher C-reactive protein (CRP) and lactate levels as well as lower platelet count when compared to the uncomplicated infection.

**Table 1.**
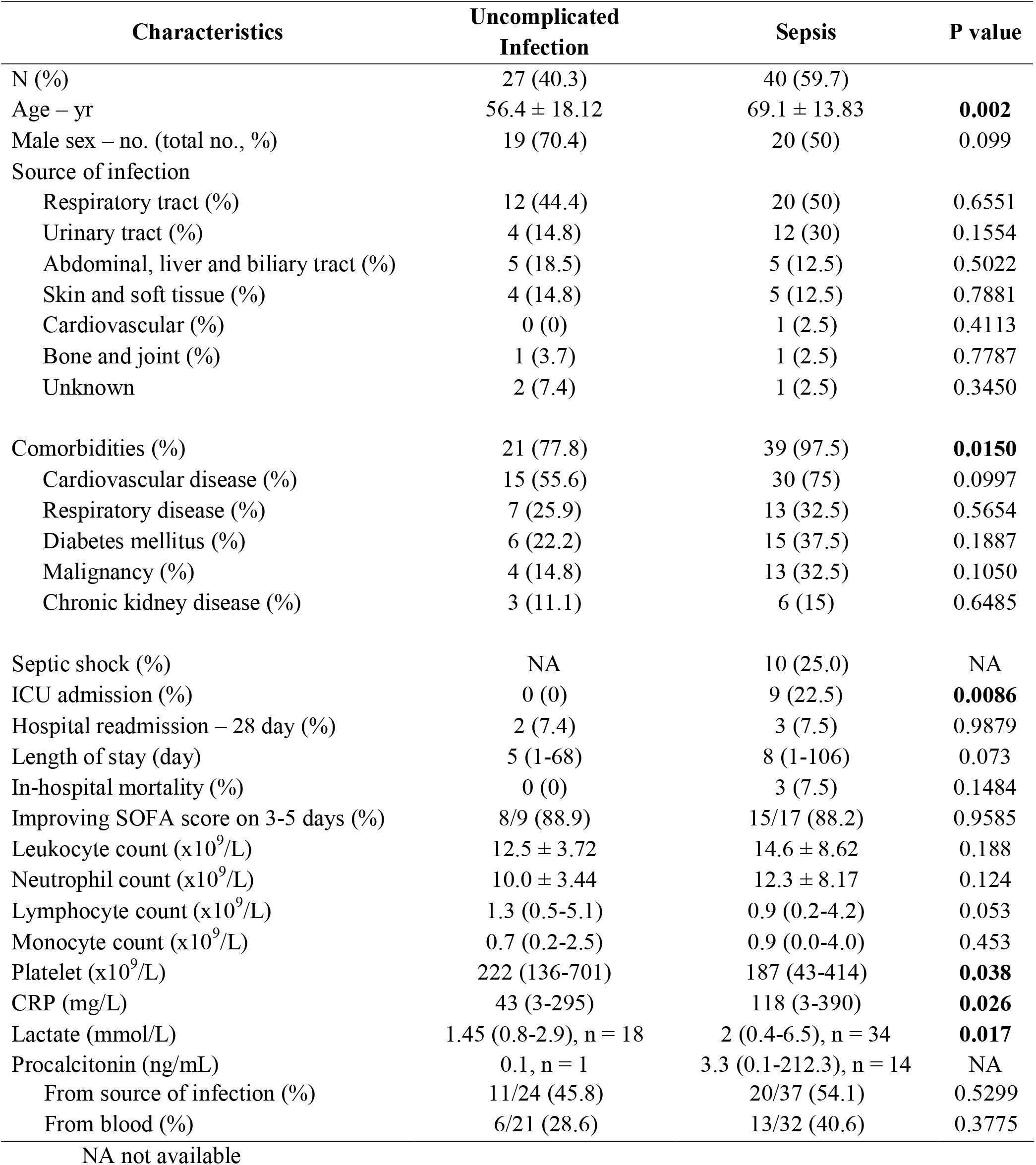
Patients Demographic and Clinical Characteristics (n = 67)

| Characteristics | Uncomplicated Infection | Sepsis | P value |
| --- | --- | --- | --- |
| N (%) | 27 (40.3) | 40 (59.7) |  |
| Age – yr | 56.4 ± 18.12 | 69.1 ± 13.83 | **0.002** |
| Male sex – no. (total no., %) | 19 (70.4) | 20 (50) | 0.099 |
| Source of infection |  |  |  |
| Respiratory tract (%) | 12 (44.4) | 20 (50) | 0.6551 |
| Urinary tract (%) | 4 (14.8) | 12 (30) | 0.1554 |
| Abdominal, liver and biliary tract (%) | 5 (18.5) | 5 (12.5) | 0.5022 |
| Skin and soft tissue (%) | 4 (14.8) | 5 (12.5) | 0.7881 |
| Cardiovascular (%) | 0 (0) | 1 (2.5) | 0.4113 |
| Bone and joint (%) | 1 (3.7) | 1 (2.5) | 0.7787 |
| Unknown | 2 (7.4) | 1 (2.5) | 0.3450 |
| Comorbidities (%) | 21 (77.8) | 39 (97.5) | **0.0150** |
| Cardiovascular disease (%) | 15 (55.6) | 30 (75) | 0.0997 |
| Respiratory disease (%) | 7 (25.9) | 13 (32.5) | 0.5654 |
| Diabetes mellitus (%) | 6 (22.2) | 15 (37.5) | 0.1887 |
| Malignancy (%) | 4 (14.8) | 13 (32.5) | 0.1050 |
| Chronic kidney disease (%) | 3 (11.1) | 6 (15) | 0.6485 |
| Septic shock (%) | NA | 10 (25.0) | NA |
| ICU admission (%) | 0 (0) | 9 (22.5) | **0.0086** |
| Hospital readmission – 28 day (%) | 2 (7.4) | 3 (7.5) | 0.9879 |
| Length of stay (day) | 5 (1-68) | 8 (1-106) | 0.073 |
| In-hospital mortality (%) | 0 (0) | 3 (7.5) | 0.1484 |
| Improving SOFA score on 3-5 days (%) | 8/9 (88.9) | 15/17 (88.2) | 0.9585 |
| Leukocyte count (x10<sup>9</sup>/L) | 12.5 ± 3.72 | 14.6 ± 8.62 | 0.188 |
| Neutrophil count (x10<sup>9</sup>/L) | 10.0 ± 3.44 | 12.3 ± 8.17 | 0.124 |
| Lymphocyte count (x10<sup>9</sup>/L) | 1.3 (0.5-5.1) | 0.9 (0.2-4.2) | 0.053 |
| Monocyte count (x10<sup>9</sup>/L) | 0.7 (0.2-2.5) | 0.9 (0.0-4.0) | 0.453 |
| Platelet (x10<sup>9</sup>/L) | 222 (136-701) | 187 (43-414) | **0.038** |
| CRP (mg/L) | 43 (3-295) | 118 (3-390) | **0.026** |
| Lactate (mmol/L) | 1.45 (0.8-2.9), n = 18 | 2 (0.4-6.5), n = 34 | **0.017** |
| Procalcitonin (ng/mL) | 0.1, n = 1 | 3.3 (0.1-212.3), n = 14 | NA |
| Positive culture |  |  |  |
| From source of infection (%) | 11/24 (45.8) | 20/37 (54.1) | 0.5299 |
| From blood (%) | 6/21 (28.6) | 13/32 (40.6) | 0.3775 |

NA not available

### Serum cytokines’ level are not significantly different between uncomplicated infection and sepsis patients

Infection/ sepsis was associated with changes in immune cell functions. We therefore sought to determine if the levels of serum cytokines can reflect changes in immune cell functions during the development of sepsis. In keeping with current literature (20), several inflammatory cytokines were higher in both uncomplicated infection and sepsis groups when compared to healthy controls (Fig 2). Serum levels of IL-10 and TNF-α were significantly higher in sepsis group than healthy controls but no difference was observed between sepsis and uncomplicated infection. Similarly, IL-6 level was higher in uncomplicated infection/ sepsis patients compared to healthy controls but again no difference was observed between sepsis and uncomplicated infection.

**Fig 1.**
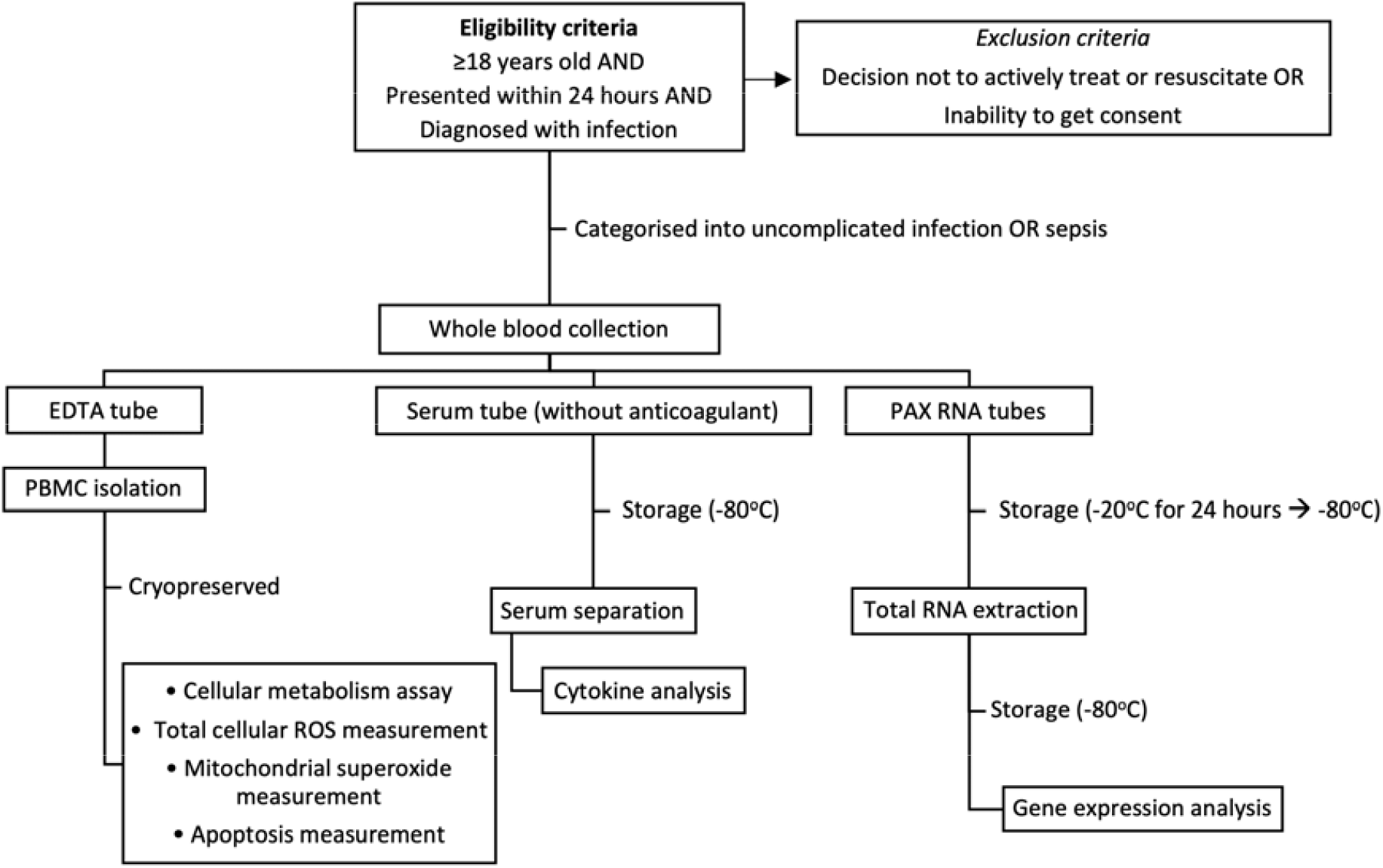
illustrates the workflow of experiment.

**Fig 2.**
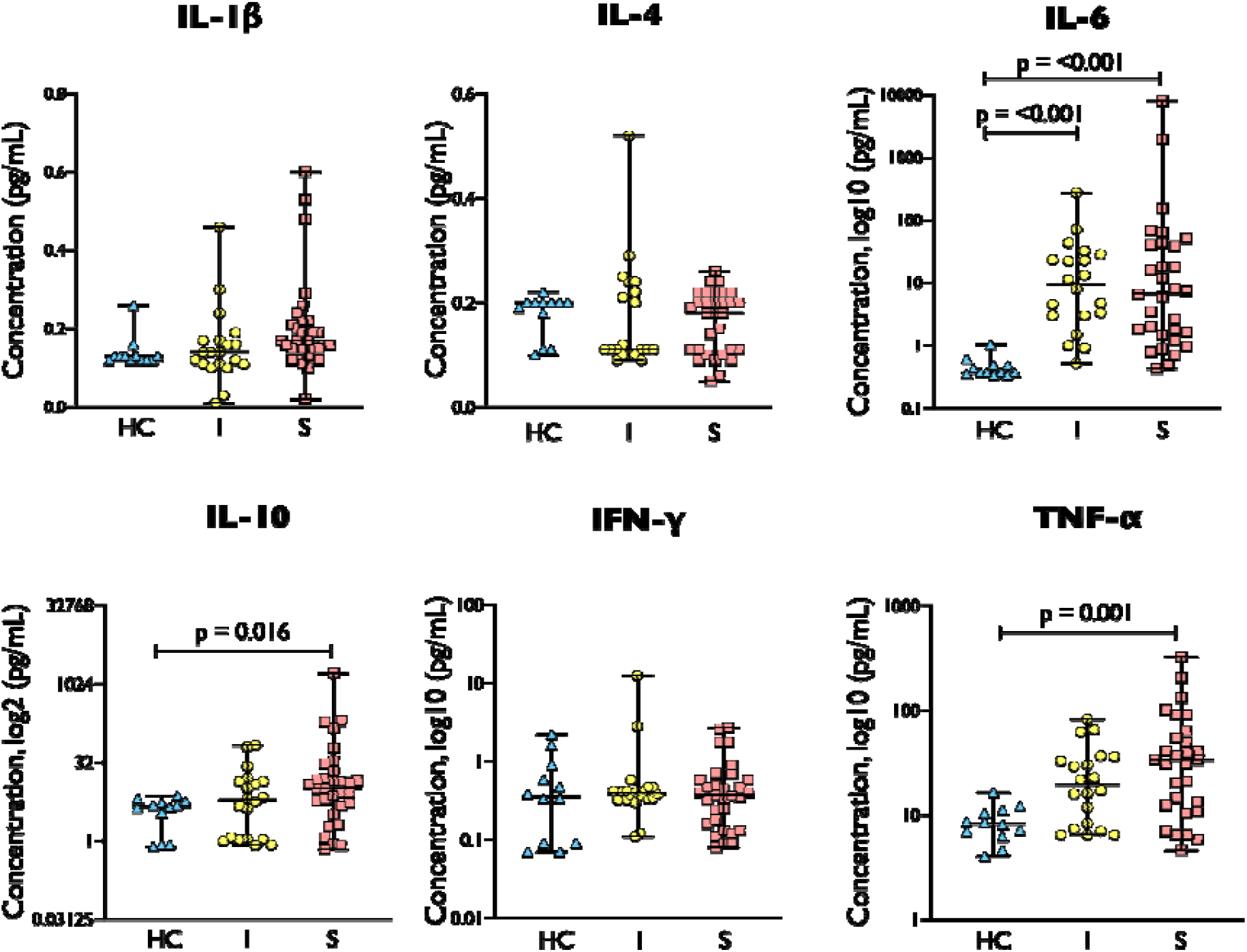
Cytokine levels in healthy control/ HC (n = 12), uncomplicated infection/ I (n = 20) and sepsis/ S (n = 31). Sepsis demonstrated the highest cytokine levels in IL-6, IL-10 and TNF-α. Comparison between groups were performed by Kruskal-Wallis test followed by Dunn’s multiple comparison test.

### Downregulations of mitochondrial function-related genes are more prominent in sepsis patients

Having found that inflammatory cytokines did not distinguish between uncomplicated infection and sepsis, we next proceeded to investigate whether gene-expression profiling could distinguish between such patients. As an initial explorative step, we performed gene expression profiling in the first 29 patients, as well as 11 healthy controls, recruited into the study (Supplementary results, S1-S2 Tables).

In these experiments, 29 out of a total of 90 genes showed differential expression (FDR <0.05) across three groups – healthy controls, uncomplicated infection and sepsis groups (Figure 3A). From these 29 genes, 17 genes were significantly downregulated in infected/ sepsis patients when compared to healthy controls (Figure 3B). Of these 17 genes, 13 genes were downregulated in the sepsis group but not in the uncomplicated infection groups. The remaining 4 genes were also downregulated in the uncomplicated infection. Twelve genes showed upregulation in infected/ sepsis patients compared to healthy controls (Figure 3B). Of those genes, 5 genes were upregulated in uncomplicated infection as well as in sepsis group. The other 7 genes were upregulated only in sepsis group.

**Fig 3A.**
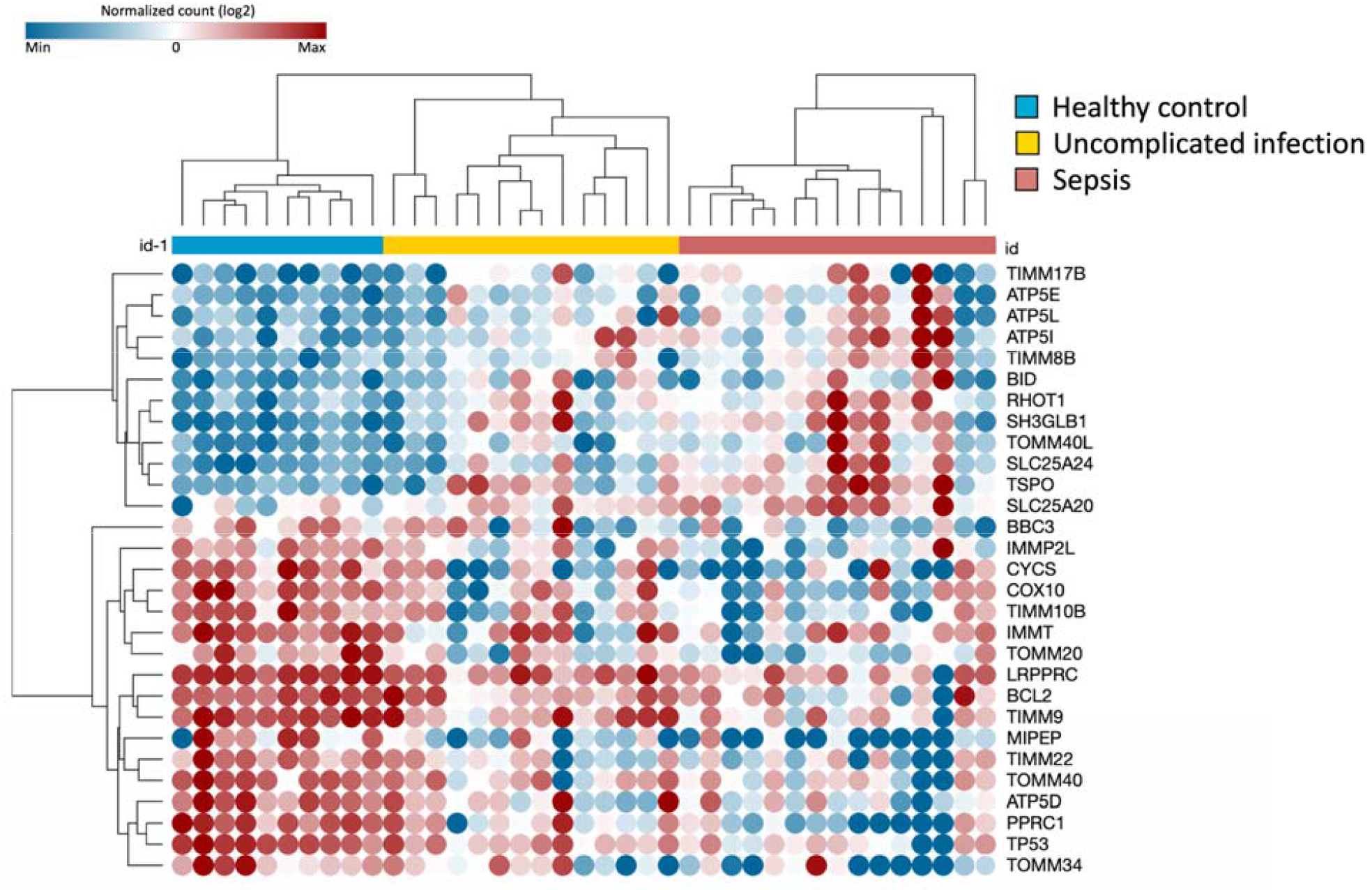
Heat map demonstrated 29 differentially expressed genes in log2-counts. The samples were grouped using semi-supervised clustering. Each sample is labelled by colour according to the clinical condition (performed on a subset of 10 healthy controls, 14 uncomplicated infection and 15 sepsis subjects).

**Fig 3B.**
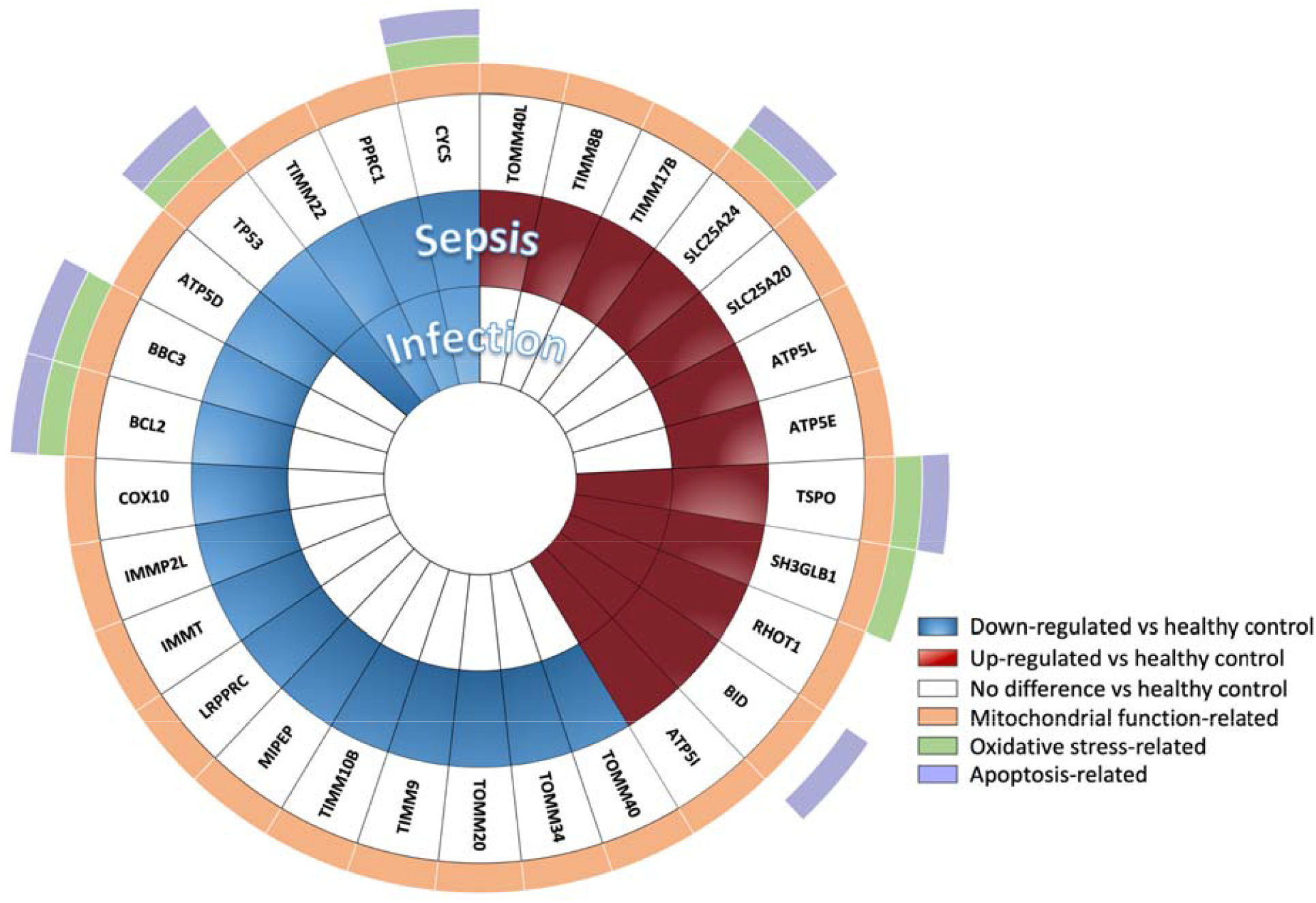
The differentially expressed genes (FDR <0.05) in uncomplicated infection (n = 14) and sepsis patients (n = 15) relative to healthy controls’ (n = 10). Function of each gene is indicated by colour (e.g. regulations on mitochondrial function, ROS production and apoptosis cell death). Twenty genes were expressed differently in sepsis. Comparison between groups were performed by one-way ANOVA followed by Benjamini-Yakutieli False Discovery Rate method.

### Impaired mitochondrial function is observed in uncomplicated infection and sepsis

Having discovered the differences in gene expression related to mitochondrial pathway between uncomplicated infection and sepsis, we were to confirm it by measuring mitochondrial function of PBMC. Parameters related to mitochondrial functions including basal respiration, maximal respiration, spare respiratory capacity and ATP production reduced significantly in sepsis when compared to healthy control group (Figure 4 A-D). When the sepsis was compared to uncomplicated infection, maximal respiration, spare respiratory capacity and ATP production were significantly lower in sepsis group. Besides mitochondrial dysfunction, we also observed a reduced basal ECAR in sepsis when compared to healthy control (Figure 4E).

**Fig 4.**
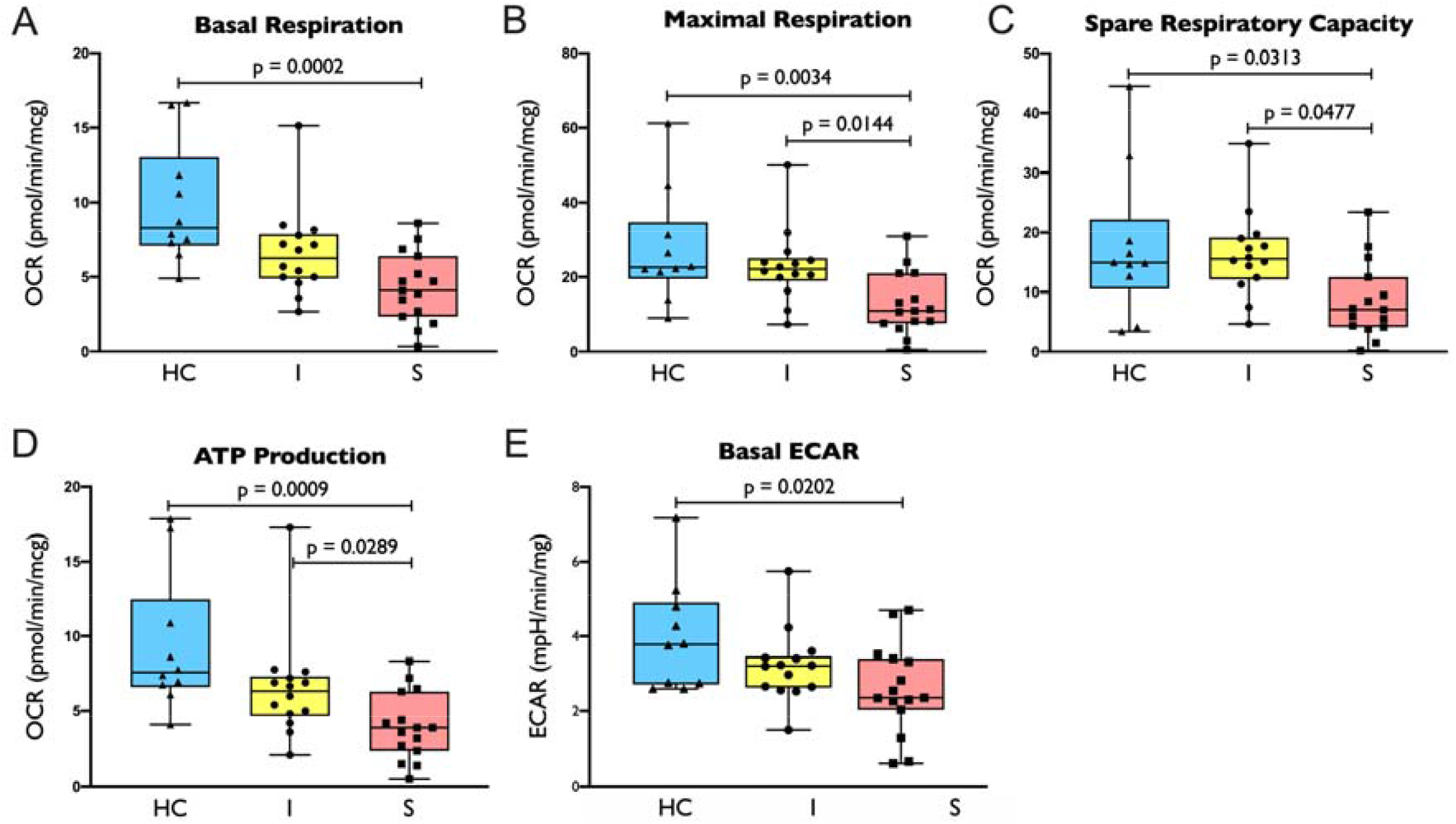
Oxygen consumption rate (OCR) and extracellular acidification rate (ECAR) on subjects with gene expression data: 4A. basal respiration, 4B. maximal respiration, 4C. spare capacity, 4D. ATP production, 4E. basal ECAR in healthy control/ HC (n = 10), uncomplicated infection/ I (n = 14) and sepsis/ S (n = 15) group. Comparison between groups were performed by one-way ANOVA followed by Tukey’s multiple comparison test.

Notably, there were 20 mitochondrial-related genes which were differentially expressed in those with sepsis but not in uncomplicated infection patients (Figure 3B). This finding points to a potential difference in leukocyte biology between uncomplicated infection and sepsis.

Having confirmed the evidence of impaired cellular metabolism in the above 29 patients, we next proceeded to measure the same metabolic parameters for the entire cohort (67 patients and 20 controls). These measurements again confirmed significant impairment and a trend towards impaired mitochondrial functions in sepsis and uncomplicated infection patients, respectively (Supplementary results, S1 Fig). The following findings described hereafter were performed on the entire cohort.

### Increased mitochondrial oxidative stress is observed in uncomplicated infection and sepsis patients

Previous research shows that increased mitochondrial oxidative stress is associated with impaired mitochondrial function in circulating leukocytes (21). Our results support this, as we observed increased mitochondrial superoxide level, as measured by MitoSOX, in both uncomplicated infection and sepsis groups when compared to healthy controls (Figure 5A). There was also a trend towards increased total cellular ROS (measured by DCFDA) in uncomplicated infection and sepsis when compared to healthy controls (Figure 5B) even though they were not statistically significant.

**Fig 5.**
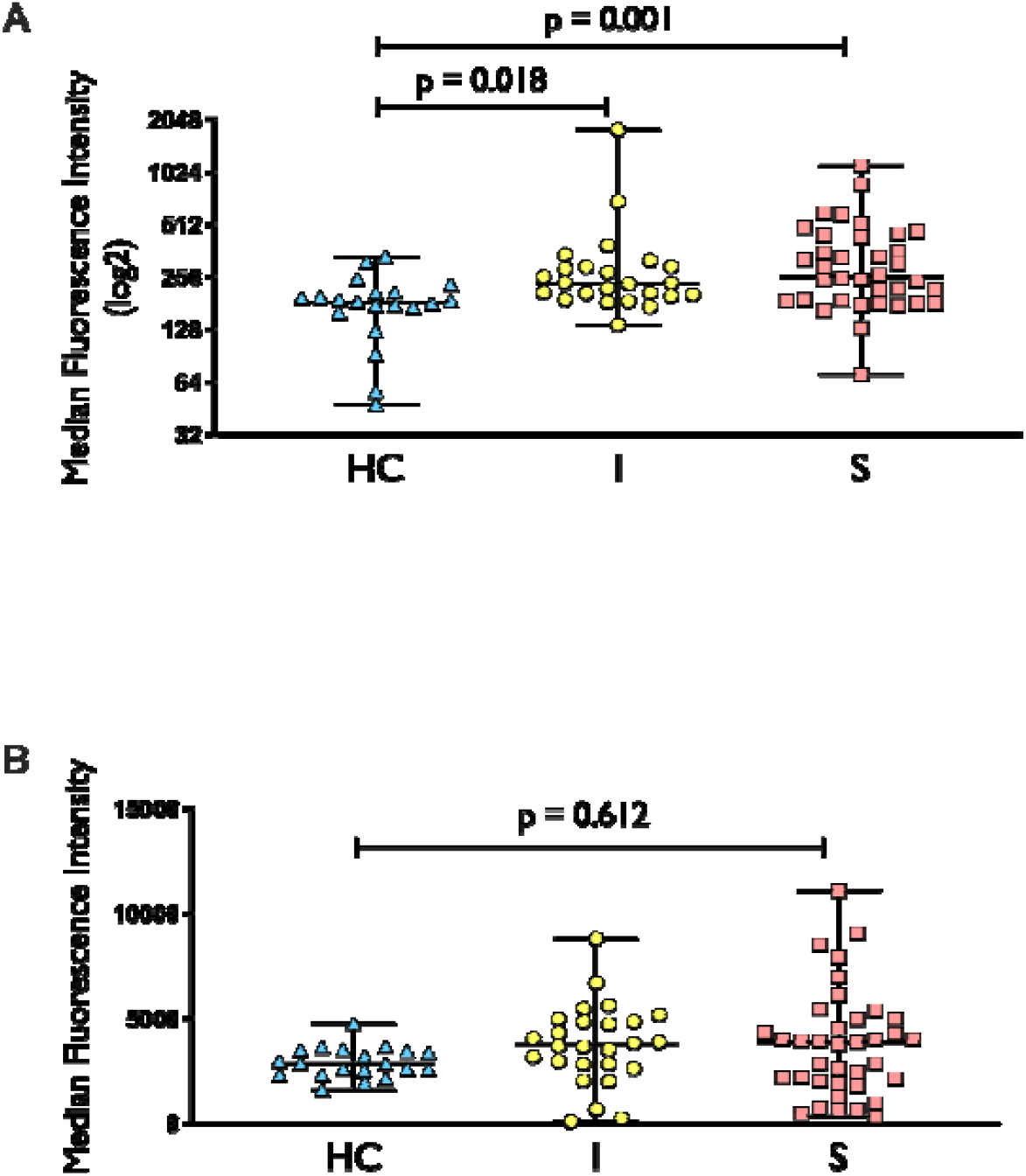
MitoSOX (5A) and DCFDA (5B) staining in healthy control (HC), uncomplicated infection (I) and sepsis (S). N for MitoSOX = HC (20), I (24), S (36), respectively; n for DCFDA = HC (20), I (26), S (37), respectively. For MitoSOX dataset, comparison between groups was performed with Kruskal-Wallis test followed by Dunn’s multiple comparison. For DCFDA dataset, comparison between groups was made using one-way ANOVA followed by Tukey’s multiple comparison.

### Mitochondrial oxidative stress correlates with gene expression level, serum cytokine level and bacteremia

Previous study showed that mitochondrial oxidative stress may be increased by several factors in infection/ sepsis (22). These factors include increased systemic inflammation (e.g. proinflammatory cytokines), pathogen factor (presence of bacteremia) and transcriptional factor (gene expression). To investigate the possible roles of those factors, we performed a correlation analysis between mitochondrial superoxide levels and: serum cytokine levels, bacteremia and mitochondrial ROS-related genes including Cytochrome C/ CYCS (electron transport chain), Tumor Protein P53/ TP53 (maintains cellular redox status), Solute Carrier Family 25 Member 24/ SLC25A24 (buffers calcium level in mitochondrial matrix) and Translocator Protein/ TSPO (translocator protein for release of proapoptotic mitochondrial component).

Our analyses showed that mitochondrial superoxide level correlated with increased proinflammatory cytokine TNF-α (*r* = 0.330, p <0.05) but not with IL-6 and IL-10 (p >0.05). Similarly, mitochondrial superoxide level correlated with expression levels of CYCS, TP53, SLC24A24 and TSPO (*r*_s_ = −0.4926, −0.4422, 0.4382, 0.4835, respectively; p <0.05). Additionally, a higher oxidative stress level was observed in patients with as compared to those without bacteremia (p <0.05) (S2 Fig).

### Impact of increased oxidative stress on leukocyte function and survival

With increased levels of mitochondrial superoxide observed in both uncomplicated infection and sepsis groups, we evaluated the association of this increase with leukocyte mitochondrial functions. Our analyses revealed a significant inverse correlation between an increased mitochondrial superoxide level and mitochondrial function (p <0.05) (S3 Fig). The findings were consistent across all parameters as measured by our cellular metabolism assay.

Existing literature shows both impaired mitochondrial function (23) and increased intra-mitochondrial oxidative stress (24) may lead to the activation of apoptotic pathways. However, we did not observe any significant increase in apoptosis measured by Annexin V and PI in PBMCs from either the uncomplicated infection or the sepsis group (S4 Fig).

## Discussion

Currently, it remains unclear whether defective cellular metabolism exists in circulating leukocytes of patients who are in the early stage sepsis. Our study provide evidence that the defect is present in patients who are infected and more so in those who progress to sepsis. The impaired cellular metabolism is evident across several platforms including gene-expression profiling, cellular metabolism analyses and intra-mitochondrial reactive oxygen species measurement.

Although we do not have direct evidence to implicate oxidative stress as a causal factor in the impairment of cellular metabolism, we do observe a moderate correlation between the degree of mitochondrial oxidative stress and the extent of cellular respiration impairment, i.e. increasing mitochondrial oxidative stress correlates with worsening of cellular respiration. The increase in mitochondrial oxidative stress level is associated with higher proinflammatory cytokine, bacteremia and gene dysregulation. Inflammatory cytokine (25) and dysregulation of genes (26) are known to be associated with oxidative stress and, in general, reduced cellular respiration (27). The presence of bacteremia, as shown in this study, is also associated with increased oxidative stress since bacterial infection is the trigger of ROS production (28).

The relationship between proinflammatory cytokine, oxidative stress and impaired mitochondrial function is complex. The levels of proinflammatory cytokines significantly show correlation with oxidative stress and impaired mitochondrial function. Mitochondrial oxidative stress itself can further activate inflammatory response through the production of signalling molecules that propagate cellular dysfunction (29). Therefore, it is likely that changes in systemic inflammation milieu, intra-mitochondrial oxidative stress and cellular respiration are intricately linked. It is beyond the scope of our study to delineate such complex relationship; clearly, additional mechanistic studies are needed in the future.

We also observe that in uncomplicated infection patients, the circulating immune cells do not shift the ATP production to glycolysis. Existing literatures indicate that host cells expect to switch their main metabolism into glycolysis when stressed (8, 30). The lack of such a switch might be because immune cells may utilize pathways alternative to glycolysis (31) to generate more ATP. Additionally, different immune cell subsets prefer to use different dominant type of metabolism, e.g. naïve T cells are dependent on mitochondrial respiration as their primary metabolic pathway while activated T cells exhibit higher glycolysis (32). Further studies are needed to address this issue, as by using glycolysis-specific assay in purified cell subsets.

This study demonstrates the lack of cytokine use as a marker for staging sepsis patients (33). The increase of cytokines found in this study could not distinguish sepsis from the uncomplicated infection, on which the routine laboratories, such as CRP, lactate level and platelet count, could discriminate those severities better. This indicates the need to develop more specific yet practical markers that enable us to stage infection patients according to their risk severities.

Our findings have some important therapeutic implications. The observed increased mitochondrial oxidative stress in uncomplicated infection could contribute to later mitochondrial dysfunction. This observation should be stretched in future mechanistic studies. If the relationship between mitochondrial oxidative stress and mitochondrial impairment is confirmed, it may open up opportunity for early intervention (e.g. mitochondrial-targeted antioxidant) for patients in early stage of infection.

We have used multiple modalities from transcriptomic to functional analyses to investigate the change in mitochondrial and immune function which has strengthened this research. Additionally, this study included infection patients from ED instead of the “established” sepsis from intensive care unit (ICU). Therefore, we could observe metabolic changes in broader spectrum of disease rather than simply comparing the changes between severe disease and the normal end. In regard to it, the finding of impaired cellular metabolism in uncomplicated infection consequently suggests that past biomarkers developed in the critically ill, i.e. ICU, patients might have low relevance in ED setting.

Some limitations are notified in this study. First, the study participants are highly heterogenous in comorbidities and age. Those factors should be put into consideration when interpreting the result, i.e. patients with diabetes mellitus or older age might have different basal oxidative stress level. Secondly, most of the study participants have favorable outcome, i.e. minimal number of mortality. In the same manner, the analysis is ideally explored in patients with poor outcome, as the result might be variable from those with good prognosis. Thirdly, total ROS is not an ideal assay to measure oxidative stress since, technically, intermediate radical will falsely amplify the fluorescence intensity (34, 35). The non-significant signal of total cellular ROS might be explained by this issue.

Surprisingly, we did not observe increased cell death in this study, despite significant increase in mitochondrial oxidative stress levels and impairment of mitochondrial function. Similar findings are also noted in previous studies (36, 37) who showed no, or slight, increase in apoptosis from flow cytometry measurement in sepsis patients. This could be because dying cells are rapidly removed by reticuloendothelial system and, therefore, are less likely to appear in circulating blood (38). Another possible explanation could be that dying cells are not captured during sample processing procedure. To address this issue, early marker of apoptosis can be used in future study (39, 40).

In conclusion, impairment of cellular metabolism, presented mainly as mitochondrial dysfunction, is evident in the immune cells of infection patients without presenting sepsis. Oxidative stress, along with inflammation and transcription regulation, might be a potential initiator, amplifier or victim to explain the impairment.

## Supporting information

Supplementary results

## Data Availability

The data referred to in the manuscript will be available from the corresponding author upon reasonable request.

## Acknowledgement

We gratefully acknowledge the contribution of Joey Lai, PhD, Stephen Schibeci, Nicole Fewings, PhD and Lawrence Ong, PhD for their input in data analysis; Sydney Informatics Hub, a Core Research Facility of the University of Sydney; and Indonesia Endowment Fund for Education (LPDP) for the grant support. No compensation was provided for those assistances.

## Supporting information

### Supplementary results

Downregulation of mitochondrial function-related genes is observed in sepsis Impaired mitochondrial function is observed in uncomplicated infection and sepsis

### Supplementary tables

**S1 Table**. List of genes on NanoString’s mitochondrial biogenesis and function panel

**S2 Table**. Demographic and clinical characteristic of overall subjects vs subset of subjects for gene expression analysis

### Supplementary figures

**S1 Fig**. Oxygen consumption rate (OCR) and extracellular acidification rate (ECAR) on subjects with gene expression data

**S2 Fig**. MitoSOX level in non-bacteremia and bacteremia subjects

**S3 Fig**. Correlation matrix between MitoSOX level and mito stress test parameters

**S4 Fig**. Annexin V^+^ and propidium iodide^-^ population in healthy control, uncomplicated infection and sepsis

