## Supplementary results for "Impaired peripheral mononuclear cell metabolism in patients at risk of developing sepsis: A cohort study"

**Supplementary Appendix**

**Table of Content**

**Supplementary Results**

Downregulation of mitochondrial function-related genes is observed in sepsis …………..	2

Impaired mitochondrial function is observed in uncomplicated infection and sepsis ……..	2

**Supplementary Tables**

**S1 Table.** List of genes on NanoString’s mitochondrial biogenesis and function panel ……………………………………………………………………………………………… 3

**S2 Table.** Demographic and clinical characteristic of overall subjects vs subset of

subjects for gene expression analysis ………………………………………………………	4

**Supplementary Figures**

**S1 Fig.** Oxygen consumption rate (OCR) and extracellular acidification rate (ECAR)

on subjects with gene expression data ……………………………………………………..	5

**S2 Fig.** MitoSOX level in non-bacteremia and bacteremia subjects ……………………...	6

**S3 Fig.** Correlation matrix between MitoSOX level and mito stress test parameters …….	7

**S4 Fig.** Annexin V^+^ and propidium iodide^-^ population in healthy control, uncomplicated infection and sepsis ………………………………………………………………………...	8

**Supplementary Result**

**Downregulation of mitochondrial function-related genes is observed in sepsis**

We performed gene expression profiling in the first twenty-nine patients recruited into the study. We used a commercially available metabolic/mitochondria focused biomarker panel (NanoString^Ⓡ^ Technologies Inc). Ten healthy controls were also included in the gene-expression profiling experiments. Given the potential for selection bias, we compared the baseline characteristics between these selected patients against the overall cohort (n = 67). Reassuringly, we found that there were no differences between these twenty-nine patients and all other patients in the cohort in terms of age, gender, comorbidities, source of infections, microbiological results, clinical course, outcomes and laboratory findings (Table S1).

**Impaired mitochondrial function is observed in uncomplicated infection and sepsis**

Cellular metabolism was measured on the entire cohort (67 patients and 20 controls). This measurement confirmed a strong trend towards impaired mitochondrial functions across all patients. Notably, the degree of impairment was greater in the sepsis group compared to the uncomplicated infection group – a finding similar to that of the first 29 patients (Figure S1 A, 1B, 1C, 1D). However, no difference in glycolysis was found between groups as we found in the initial cohort (Figure S1 E).

**S1 Table. List of Genes on NanoString’s**

**Mitochondrial Biogenesis and Function Panel**

| AIFM2 | MIPEP | SLC25A30 | **Housekeeping** |
| --- | --- | --- | --- |
| AIP | MPV17 | SLC25A31 | ABCF1 |
| ATP5F1D | MSTO1 | SLC25A37 | GUSB |
| ATP5F1E | MTX2 | SLC25A4 | HPRT1 |
| ATP5I | NEFL | SLC25A5 | LDHA |
| ATP5L | NRF1 | SOD1 | POLR1B |
| ATP5O | OPA1 | STARD3 | RPLP0 |
| BAK1 | PMAIP1 | TAZ |  |
| BBC3 | PPRC1 | TIMM10 |  |
| BCL2 | RHOT1 | TIMM10B |  |
| BCL2L1 | RHOT2 | TIMM17A |  |
| BID | SH3GLB1 | TIMM17B |  |
| BNIP3 | SLC25A1 | TIMM22 |  |
| CDKN2A | SLC25A10 | TIMM23 |  |
| COX10 | SLC25A12 | TIMM44 |  |
| COX18 | SLC25A13 | TIMM50 |  |
| CPT1B | SLC25A14 | TIMM8A |  |
| CPT2 | SLC25A15 | TIMM8B |  |
| CYCS | SLC25A16 | TIMM9 |  |
| DNM1L | SLC25A17 | TOMM20 |  |
| FIS1 | SLC25A19 | TOMM22 |  |
| GRPEL1 | SLC25A2 | TOMM34 |  |
| IDH2 | SLC25A20 | TOMM40 |  |
| IMMP1L | SLC25A21 | TOMM40L |  |
| IMMP2L | SLC25A22 | TP53 |  |
| IMMT | SLC25A23 | TSPO |  |
| LRPPRC | SLC25A24 | UCP1 |  |
| MFN1 | SLC25A25 | UCP2 |  |
| MFN2 | SLC25A27 | UCP3 |  |
| MINOS1 | SLC25A3 | UXT |  |

**Note:** Two housekeeping genes, GUSB and RPLP0, were used to normalize the count. Another 4 housekeeping genes from this TagSet were excluded from the analyses as their expressions were found to be unstable between groups (data not shown).

**S2 Table. Demographic and Clinical Characteristic of**

**Overall Subjects vs Subset of Subjects for Gene Expression Analysis**

| **Characteristics** | **Overall** | **Subset** | **P value** |
| --- | --- | --- | --- |
| N (%) | 67 (100) | 29 (43) |  |
| Age – yr | 64.0 ± 16.76 | 62.6 ± 20.85 | 0.7248 |
| Male sex – no. (total no., %) | 39 (58) | 17 (59) | 0.9277 |
| SOFA score ≥2 (%) | 40 (59.7) | 15 (51.7) | 0.4692 |
| Source of infection |  |  |  |
| Respiratory tract (%) | 32 (47.8) | 10 (34.5) | 0.2302 |
| Urinary tract (%) | 16 (23.9) | 6 (20.7) | 0.7334 |
| Abdominal, liver and biliary tract (%) | 10 (14.9) | 7 (24.1) | 0.2805 |
| Skin and soft tissue (%) | 9 (13.4) | 6 (20.7) | 0.3608 |
| Cardiovascular (%) | 1 (1.5) | 1 (3.4) | 0.5507 |
| Bone and joint (%) | 2 (3.0) | 1 (3.4) | 0.9180 |
| Unknown | 3 (4.5) | 0 (0) | 0.2482 |
| Comorbidities |  |  |  |
| Cardiovascular disease (%) | 45 (67.2) | 21 (72.4) | 0.6156 |
| Respiratory disease (%) | 20 (29.9) | 4 (13.8) | 0.0963 |
| Diabetes mellitus (%) | 21 (31.3) | 10 (34.5) | 0.7593 |
| Malignancy (%) | 17 (25.4) | 3 (10.3) | 0.0961 |
| Chronic kidney disease (%) | 9 (13.4) | 4 (13.8) | 0.9582 |
| Septic shock (%) | 11 (16.4) | 4 (13.8) | 0.7485 |
| ICU admission (%) | 9 (13.4) | 4 (13.8) | 0.9582 |
| Hospital readmission – 28 day (%) | 5/64 (7.8) | 4/28 (14) | 0.3575 |
| Length of stay (day) | 7 (0-106) | 7 (0-106) | 0.9066 |
| In-hospital mortality (%) | 3 (4.5) | 1 (3.4) | 0.8054 |
| Improving SOFA score on 3-5 days (%) | 23/26 (88) | 6/8 (75) | 0.3758 |
| Leukocyte count (x10^9^/mm^3^) | 13.3 (1.0-37.7) | 16.0 ± 7.24 | 0.1533 |
| Neutrophil count (x10^9^/mm^3^) | 10.4 (0.0-36.1) | 13.4 ± 6.90 | 0.1378 |
| Lymphocyte count (x10^9^/mm^3^) | 1.1 (0.2-5.1) | 1.2 (0.3-4.2) | 0.6235 |
| Monocyte count (x10^9^/mm^3^) | 0.9 (0.0-4.0) | 0.9 (0.2-4.0) | 0.1725 |
| CRP (mg/L) | 102 (3-390) | 105 (3-319) | 0.6268 |
| Lactate (mmol/L) | 1.6 (0.4-6.5), n = 52 | 1.6 (0.7-6.5), n = 21 | 0.96378 |
| Procalcitonin (ng/mL) | 2.0 (0.1-212.3), n = 15 | 3.1 (0.18-212.3), n = 6 | 0.7191 |
| Positive culture |  |  |  |
| From source of infection (%) | 31/61 (51) | 10/25 (40) | 0.3566 |
| From blood (%) | 19/53 (34) | 9/25 (36) | 0.8633 |

**S1 Figure**

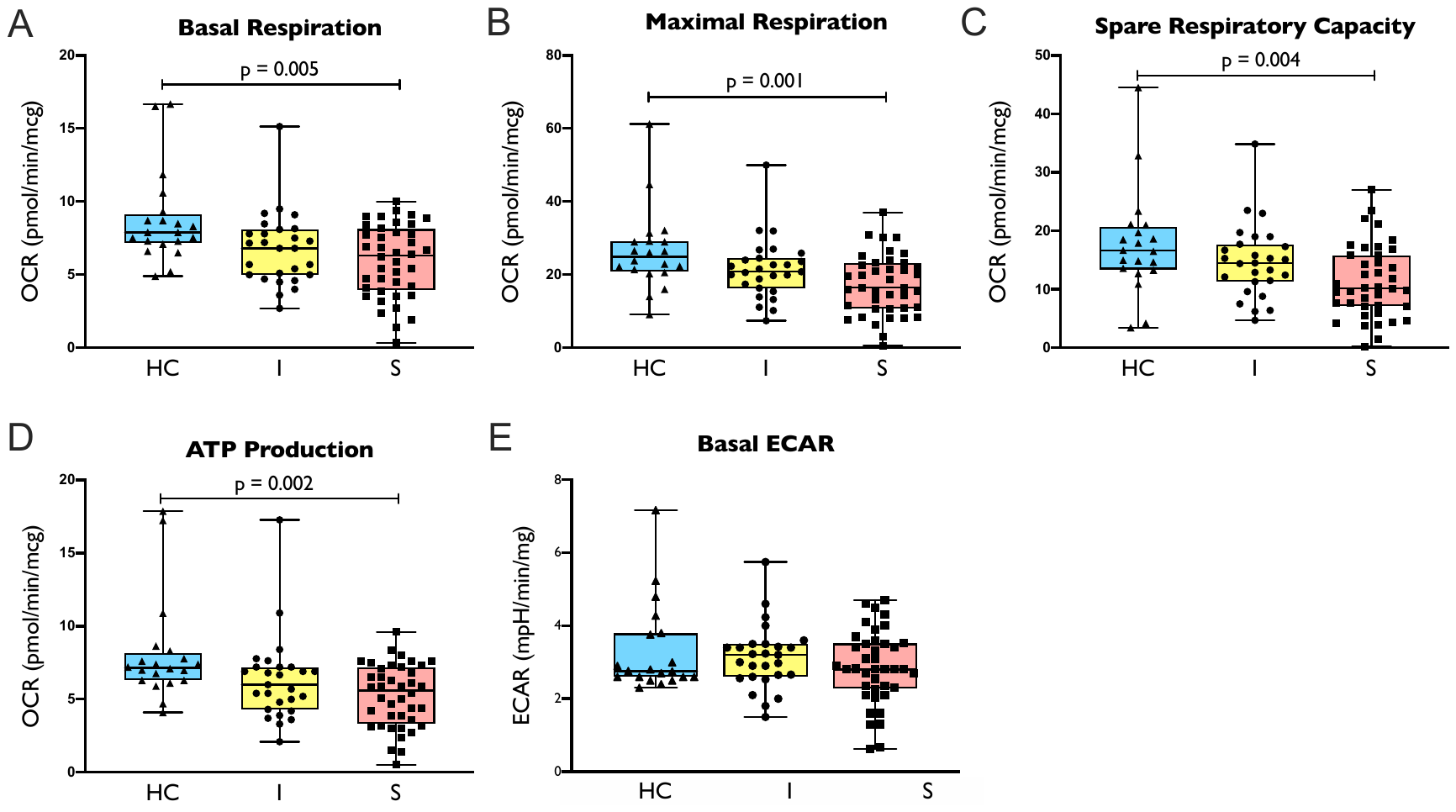

**S1 Fig.** Oxygen consumption rate (OCR) (1A. basal respiration, 1B. maximal respiration, 1C. spare capacity, and 1D. ATP production) and extracellular acidification rate (ECAR) (1E) in healthy control/ HC (n = 20), uncomplicated infection/ I (n = 27) and sepsis/ S (n = 40) group. Comparison between groups were performed by one-way ANOVA followed by Tukey’s multiple comparison test for maximal respiration and spare respiratory capacity; and Kruskal-Wallis test followed by Dunn’s multiple comparison test for basal respiration, ATP production and ECAR.

**S2 Figure**

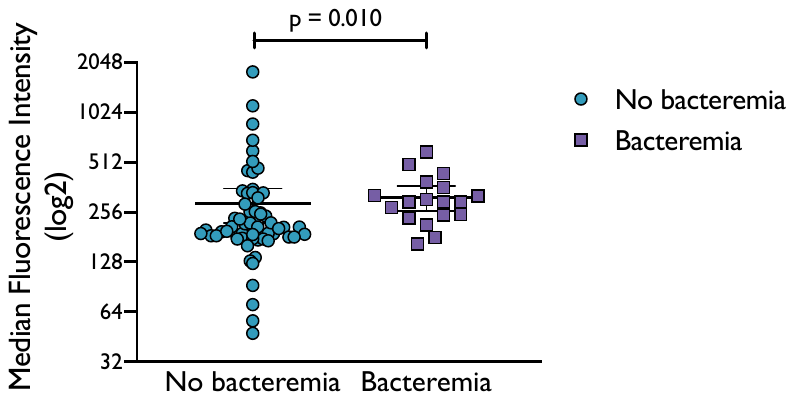

**S2 Fig.** MitoSOX level in non-bacteremia (n = 62, including healthy controls) and bacteremia (n = 18) subjects. Comparison between groups was performed with Mann-Whitney U test.

**S3 Figure**

p value (-log)

|  | Adjusted R model |
| --- | --- |
| Basal respiration | 2.30103 |
| Maximal respiration | 2.39794 |
| Spare respiratory capacity | 2.04576 |
| ATP production | 1.85387 |
| ECAR | 0.521434 |

**S3 Fig.** Correlation matrix between MitoSOX level (in Median Fluorescence Intensity) with cellular metabolism parameters (n = 80). Analysis was performed with Pearson correlation.

**S4 Figure**

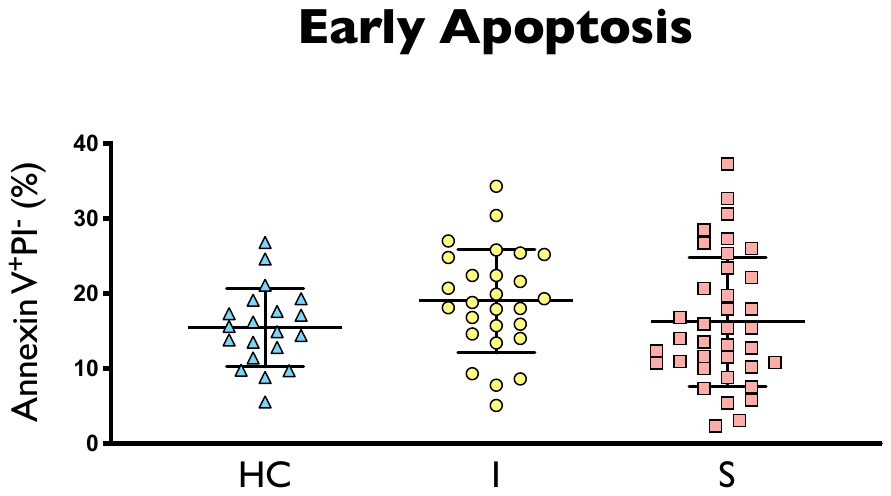

**S4 Fig.** Annexin V^+^ and propidium iodide^-^ population in healthy control (HC), uncomplicated infection (I) and sepsis (S) (n = 20, 27, 37, respectively). Comparison between groups was made using one-way ANOVA followed by Tukey’s multiple comparison.
